# Effect of Ivermectin 600 μg/kg for 6 days vs Placebo on Time to Sustained Recovery in Outpatients with Mild to Moderate COVID-19: A Randomized Clinical Trial

**DOI:** 10.1101/2022.12.15.22283488

**Authors:** Susanna Naggie, David R. Boulware, Christopher J. Lindsell, Thomas G. Stewart, Stephen C. Lim, Jonathan Cohen, David Kavtaradze, Arch P. Amon, Ahab Gabriel, Nina Gentile, G. Michael Felker, Russell L. Rothman, Dushyantha Jayaweera, Matthew W. McCarthy, Mark Sulkowski, Sybil Wilson, Allison DeLong, April Remaly, Rhonda Wilder, Sean Collins, Sarah E. Dunsmore, Stacey J. Adam, Florence Thicklin, George J. Hanna, Adit A. Ginde, Mario Castro, Kathleen McTigue, Elizabeth Shenkman, Adrian F. Hernandez, the Accelerating Covid-19 Therapeutic Interventions and Vaccines (ACTIV)-6 Study Group and Investigators

**Affiliations:** Duke Clinical Research Institute, Duke University School of Medicine, Durham, NC; Department of Medicine, Duke University School of Medicine, Durham, NC; Division of Infectious Diseases and International Medicine, University of Minnesota, Minneapolis, MN; University Medical Center, Nashville, TN; School of Data Science, University of Virginia, Charlottesville, VA; University Medical Center New Orleans, Louisiana State University Health Sciences Center, New Orleans, LA; Jadestone Clinical Research, LLC, Silver Spring, MD; David Kavtaradze, MD, Inc., Cordele, GA; Lakeland Regional Medical Center, Lakeland, FL; Focus Clinical Research Solutions, Charlotte, NC; Department of Emergency Medicine, Lewis Katz School of Medicine at Temple University, Philadelphia, PA; Department of Medicine, Miller School of Medicine, University of Miami, Miami, FL; Weill Cornell Medicine, New York, NY; Division of Infectious Diseases, Johns Hopkins University, Baltimore, MD; Veterans Affairs Tennessee Valley Healthcare System, Geriatric Research, Education and Clinical Center (GRECC), Nashville, TN; National Center for Advancing Translational Sciences, Bethesda, MD; Foundation for the National Institutes of Health, Bethesda, MD; Stakeholder Advisory Committee, Pittsburgh, PA; Biomedical Advanced Research and Development Authority, Washington, DC; University of Colorado School of Medicine, Aurora, CO; Division of Pulmonary, Critical Care and Sleep Medicine, University of Missouri-Kansas City School of Medicine, Kansas City, KS; Department of Medicine, University of Pittsburgh Medical Center, Pittsburgh, PA; Department of Health Outcomes & Biomedical Informatics, College of Medicine, University of Florida, Gainesville, FL

## Abstract

**Background:** Whether ivermectin, with a maximum targeted dose of 600 μg/kg, shortens symptom duration or prevents hospitalization among outpatients with mild to moderate coronavirus disease 2019 (COVID-19) remains unknown. Our objective was to evaluate the effectiveness of ivermectin, dosed at 600 μg/kg, daily for 6 days compared with placebo for the treatment of early mild to moderate COVID-19.

**Methods:** ACTIV-6, an ongoing, decentralized, randomized, double-blind, placebo-controlled, platform trial, was designed to evaluate repurposed therapies in outpatients with mild to moderate COVID-19. A total of 1206 participants age ≥30 years with confirmed COVID-19, experiencing ≥2 symptoms of acute infection for ≤7 days, were enrolled from February 16, 2022, through July 22, 2022, with follow-up data through November 10, 2022, at 93 sites in the US. Participants were randomized to ivermectin, with a maximum targeted dose of 600 μg/kg (n=602), daily vs. placebo daily (n=604) for 6 days. The primary outcome was time to sustained recovery, defined as at least 3 consecutive days without symptoms. The 7 secondary outcomes included a composite of hospitalization, death, or urgent/emergent care utilization by day 28.

**Results:** Among 1206 randomized participants who received study medication or placebo, median (interquartile range) age was 48 (38–58) years; 713 (59%) were women; and 1008 (84%) reported ≥2 SARS-CoV-2 vaccine doses. Median time to recovery was 11 (11–12) days in the ivermectin group and 11 (11–12) days in the placebo group. The hazard ratio (HR) (95% credible interval [CrI], posterior probability of benefit) for improvement in time to recovery was 1.02 (0.92–1.13; P[HR>1]=0.68). In those receiving ivermectin, 34 (5.7%) were hospitalized, died, or had urgent or emergency care visits compared with 36 (6.0%) receiving placebo (HR 1.0, 0.6– 1.5; P[HR<1]=0.53). In the ivermectin group, 1 participant died and 4 were hospitalized (0.8%); 2 participants (0.3%) were hospitalized in the placebo group and there were no deaths. Adverse events were uncommon in both groups.

**Conclusions:** Among outpatients with mild to moderate COVID-19, treatment with ivermectin, with a maximum targeted dose of 600 μg/kg daily for 6 days, compared with placebo did not improve time to recovery. These findings do not support the use of ivermectin in patients with mild to moderate COVID-19.

**Trial registration:** ClinicalTrials.gov Identifier: NCT04885530.

## INTRODUCTION

Despite treatment advances for coronavirus disease 2019 (COVID-19), the evolution of SARS-CoV-2 variants and subvariants has shifted therapeutic options with loss of effectiveness of monoclonal antibodies. Novel oral antivirals have been authorized for high-risk individuals in high-income countries.^1,2^ However, efficacy of these antivirals in those vaccinated or with prior infection remains unclear. Interest remains for the potential of repurposed drugs to improve symptoms and clinical outcomes in patients with COVID-19.

Numerous repurposed drugs have been investigated for COVID-19, with several large, randomized outpatient trials published.^3–5^ Trial results have been mixed. Some suggest promise to reduce emergency department (ED) visits or hospitalizations for drugs including fluvoxamine at 100 mg twice daily^3^ and immediate-release metformin.^6^ Others have failed to show a reduction in emergency department visits or hospitalizations such as fluvoxamine 50 mg twice daily.^6^ While recently completed trials benefit from increasing representation of vaccinated people, which is more relevant to the pandemic’s current state, the results have not impacted treatment guidelines due to study design and regulatory requirements.^7–9^

Ivermectin, an anti-parasitic drug used worldwide for onchocerciasis and strongyloidiasis, emerged in 2020 as a potential repurposed drug for COVID-19 informed by an *in vitro* study suggesting possible anti-viral activity.^10^ The interest for ivermectin as a COVID-19 therapeutic has remained high and, while there have been numerous ivermectin studies, its use has become controversial due to a lack of high-quality, adequately powered randomized trials and article retractions of some of the earlier and most positive studies.^11–14^ Three large randomized outpatient trials in people with symptomatic mild or moderate COVID-19 failed to identify a clinical benefit of ivermectin when dosed at 400 μg/kg daily for 3 days.^15–17^ One possibility is that the dose and duration studied was too low and too short, missing the therapeutic window for ivermectin. A proof-of-concept randomized controlled clinical trial in hospitalized patients showed that ivermectin, 600 μg/kg, for 5 days suggested possible anti-viral activity with increasing dose.^17,18^ For this reason we tested ivermectin, with a maximum targeted dose of 600 μg/kg, daily for 6 days on the Accelerating COVID-19 Therapeutic Interventions and Vaccines (ACTIV-6) platform from February 16, 2022 through July 22, 2022.

ACTIV-6 is an ongoing, fully remote (decentralized), double-blind, randomized, placebo-controlled, platform trial investigating repurposed drugs for the treatment of mild to moderate COVID-19 in the outpatient setting. This report describes the effectiveness of this dose and duration of ivermectin compared with blinded placebo for the treatment of early mild to moderate COVID-19.

## METHODS

### Trial Design and Oversight

This is a double-blind, randomized, placebo-controlled platform protocol designed to be flexible, allowing enrollment across a wide range of settings within healthcare systems, the community, and virtually. The platform enrolls outpatients with mild to moderate COVID-19 with a confirmed positive SARS-CoV-2 test. The full trial protocol and statistical analysis plan are available in **Supplement 1** and **Supplement 2**, respectively.

The protocol was approved by each site’s institutional review board. Participants provided informed consent either via written consent or an electronic consent process. An independent data monitoring committee oversaw safety and trial performance.

### Participants

Recruitment into the platform trial opened on June 11, 2021. Ivermectin 600 μg/kg was included on the platform beginning February 16, 2022. Enrollment into the ivermectin 600 μg/kg group was stopped on July 22, 2022, when 1206 participants had received their study drug, identical matched-placebo, or contributing-placebo. Participants were either identified by sites or self-identified by contacting a central study telephone hotline or website.

Study staff verified eligibility criteria including age ≥30 years, SARS-CoV-2 infection within 10 days (positive polymerase chain reaction [PCR] or antigen, including home-based test), and experiencing ≥2 symptoms of acute COVID-19 for ≤7 days from enrollment. Symptoms could include fatigue, dyspnea, fever, cough, nausea, vomiting, diarrhea, body aches, chills, headache, sore throat, nasal symptoms, and loss of sense of taste or smell. Exclusion criteria included hospitalization, study drug use within 14 days, or known allergy or contraindication to study drug (**Supplement 1**). Vaccination was allowable, as were standard of care therapies for COVID-19 available under US Food and Drug Administration (FDA) authorization or approval.

### Randomization

Participants were randomized, using a random number generator, in a 2-step process (**Figure 1**). First, participants were randomized to active agent or placebo in a ratio of *m*:1, where *m* is the number of study drugs for which the participant was eligible; the other study drug under investigation during this period was fluvoxamine, 50 mg twice daily for 10 days. Participants could choose to opt out of specific study drug groups if they or the site investigator did not feel there was equipoise, or a contraindication existed. After randomization to active agent versus placebo, participants were randomized with equal probability among the study drugs for which they were eligible. The more study drugs a participant was eligible for, the greater the chance of receiving an active agent. Participants eligible for both ivermectin and fluvoxamine 50 mg but randomized to fluvoxamine-matched placebo were included and *contributed* to the placebo group for ivermectin.

**Figure 1.**
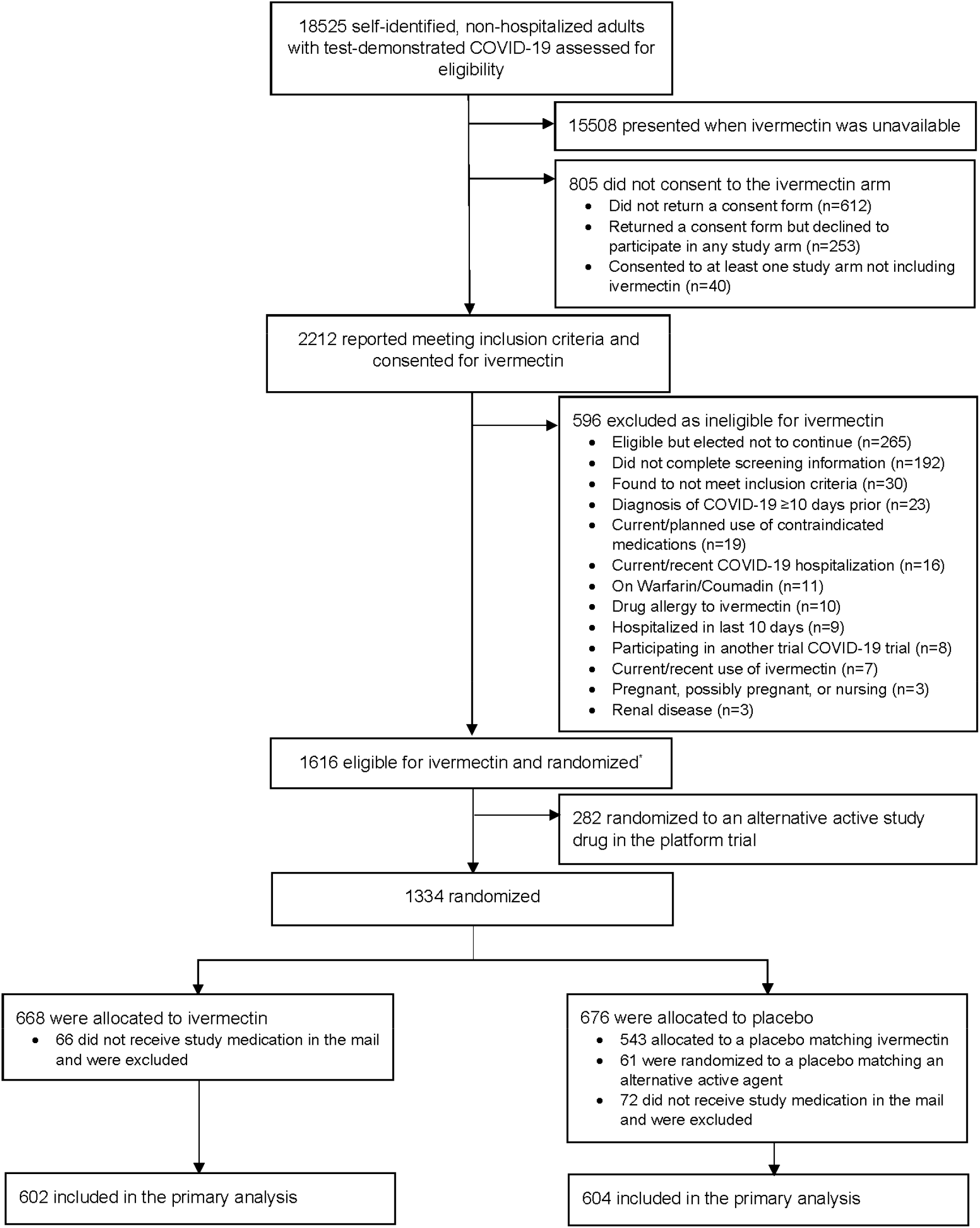
CONSORT diagram

### Interventions

A central pharmacy supplied ivermectin or placebo to participants via direct home delivery. Ivermectin was supplied as a bottle of 7-mg tablets. Participants were instructed to take a pre-specified number of tablets for 6 consecutive days based on their weight for a maximum daily dose of approximately 600 μg/kg. The dosing schedule was based on weight (kg) ranges as follows: 35-52, 53-69, 70-89, 90-109, 110-129, and >129. This schedule resulted in a range of doses from 400-600 μg/kg (**eFigure 1**). Packaging for matched placebo was identical to ivermectin while packaging for the contributing placebos was identical to that of the associated study drug, which in this case was fluvoxamine 50mg.

### Outcome Measures

The primary measure of effectiveness was time to sustained recovery, defined as the number of days between study drug receipt and the third of 3 consecutive days without symptoms. This was selected *a priori* from among the 2 co-primary endpoints that remain available to other study drugs in the platform (**Supplement 2**). The key secondary outcome was the composite of hospitalization or death by day 28. Other secondary outcomes included mean time unwell estimated from a longitudinal ordinal model; COVID-19 Clinical Progression Scale on days 7, 14, and 28; mortality through day 28; and the composite of urgent or emergency care visit, hospitalization, or death through day 28.

### Trial Procedures

The study was designed as a fully remote, or decentralized, trial. Screening and eligibility confirmation were participant-reported and site confirmed. A positive SARS-CoV-2 PCR or antigen test result was verified prior to randomization. At screening, participant-reported demographic information was collected and included race and ethnicity, eligibility criteria, medical history, concomitant medications, symptom reporting, and quality of life questionnaires.

A central investigational pharmacy distributed study drug (either active or placebo) using a next-day priority shipping service. Delivery was tracked, delayed deliveries followed-up on, and participants must have received study drug within 7 days of enrollment to be included. Confirmation that the study drug was delivered to the participant’s address was required for the participant to be included in the analysis. Receipt of study drug was defined as study day 1.

We asked participants to complete daily assessments and report adverse events through day 14. Assessments included symptoms and severity, health care visits, and medications. If symptoms were still ongoing at day 14, daily surveys continued until participants experienced 3 consecutive days without symptoms or until day 28. At days 28 and 90, all participants completed assessments. **Supplement 1** presents survey details. Additional details are available in **Supplement 3**.

### Statistical Analysis Plan

This platform trial was designed to be analyzed accepting the possibility of adding and dropping groups as the trial progresses. The general analytical approach was regression modelling. We utilized proportional hazard regression for time-to-event analysis and cumulative probability ordinal regression models for ordinal outcomes. In addition, we estimated mean time spent unwell using a longitudinal ordinal regression model as a quantification of benefit (**Supplement 2**).

The planned primary endpoint analysis was a Bayesian proportional hazards model for time to sustained recovery. The primary inferential (decision-making) quantity was the posterior distribution for the treatment assignment hazard ratio (HR), with HR>1 indicating faster recovery. If at any of the interim or final analyses, the posterior probability of benefit exceeded 0.95 (i.e. ≥95% probability of benefit), the trial would conclude efficacy of ivermectin. To preserve type 1 error <0.05, the prior for the treatment effect parameter (on the log_e_ relative hazard scale) was a normal distribution centered at 0 and scaled to a standard deviation of 0.1. All other parameter priors were non-informative, using the software default of 2.5 times the ratio of the standard deviation of the outcome divided by the standard deviation of the predictor variable. The study design was estimated to have 80% power to detect a HR of 1.2 in the primary endpoint with approximately 1200 participants. To achieve this sample size in an ongoing platform trial, once 1200 participants had been randomized to the study group or matching placebo and had received study drug, the study arm became unavailable for new participants expressing interest in the platform. Some participants had already consented to participate but had not yet been randomized or received study drug at the time of arm closure, and these participants were allowed to continue as assigned.

The primary endpoint adjusted model included the following predictor variables in addition to randomization assignment: age (as restricted cubic spline), sex, duration of symptoms at study drug receipt, calendar time (as restricted cubic spline), vaccination status, geographic region (Northeast, Midwest, South, West), call center enrollment, and day 1 symptom severity.

This adjusted model was pre-specified. The proportional hazards assumption of the primary endpoint was evaluated by generating visual diagnostics such as the log-log plot and plots of time-dependent regression coefficients for each predictor in the model, a diagnostic which indicates deviations from proportionality if the time-dependent coefficients are not constant in time.

Secondary endpoints were analyzed with Bayesian regression models (either proportional hazards or proportional odds) using non-informative priors for all parameters. Secondary endpoints were not used for formal decision making, and no decision threshold was selected. Due to potential for inflated type 1 error due to multiple comparisons, secondary endpoints should be interpreted as exploratory. The same covariates used in the primary endpoint model were used in the adjusted analysis of secondary endpoints, provided the endpoint accrued enough events to be analyzed with covariate adjustment.

As a platform trial, the primary analysis is implemented separately for each study drug, where the placebo group consists of contemporaneously randomized participants who met the eligibility criteria for that study drug; this includes both matched and contributing placebo. From other remote trials,^3,19^ we recognized that study drug delivery may not always occur (e.g., failure of delivery, participant withdrawal, or interval hospitalization). For ACTIV-6, the modified-intent-to-treat (mITT) analysis set for the primary analyses included all participants who received study drug, and participants were analyzed as assigned. We used all available data to compare ivermectin versus placebo, regardless of post-randomization adherence. In both the primary and secondary endpoint analyses, missing data among covariates used for adjustment was addressed with conditional mean imputation as the amount of missing covariate data was minimal (<4%).

A pre-specified analysis tested for differential treatment effects as a function of pre-existing participant characteristics. Analysis of heterogeneity of treatment effect included: age, symptom duration, body mass index, day 1 symptom severity, calendar time (surrogate for SARS-CoV-2 variant), sex, and vaccination status; continuous variables were modelled as such without creating subgroups.

Analyses were performed with R 4.1 with primary packages of: rstanarm, rmsb, and survival.^20^

Additional details are available in **Supplement 3**.

## RESULTS

### Study Population

Of the 2212 participants who consented to be evaluated for inclusion in the ivermectin group, 1206 were eligible for ivermectin, randomized to ivermectin, with a targeted maximum dose of 600 μg/kg, (n=602) or placebo (n=604); and received study drug (**Figure 1**). Of participants receiving placebo, 543 (90%) received matching placebo and 61 (10%) received placebo as part of the contributing placebo group.

The median age of the participants was 48 years (IQR 38–58), and 46% were aged 50 years or older (**Table 1**). The population was 59% female, 7.7% identified as Black/African American, 75% identified as White, and 21.6% reported being of Latino/Hispanic ethnicity. Although not required for enrollment, high-risk comorbidities included body mass index >30 kg/m^2^ (38%), diabetes (9%), hypertension (27%), asthma (14%), and chronic obstructive pulmonary disease (2%). Overall, 84% of participants reported receiving ≥2 COVID-19 vaccine doses. Median time from symptom onset to enrollment was 3 days (IQR 2–5) and to study drug receipt was 5 days (IQR 3–7) with 60% receiving study drug within 5 days of symptom onset (**eFigure 2**). **eTable 1** presents baseline symptom prevalence and severity. Receipt of FDA-authorized therapies was uncommon (remdesivir 0%, monoclonal antibody 3.9%, nirmatrelvir/ritonavir 3.4%, molnupiravir 0.5%).

**Table 1.**
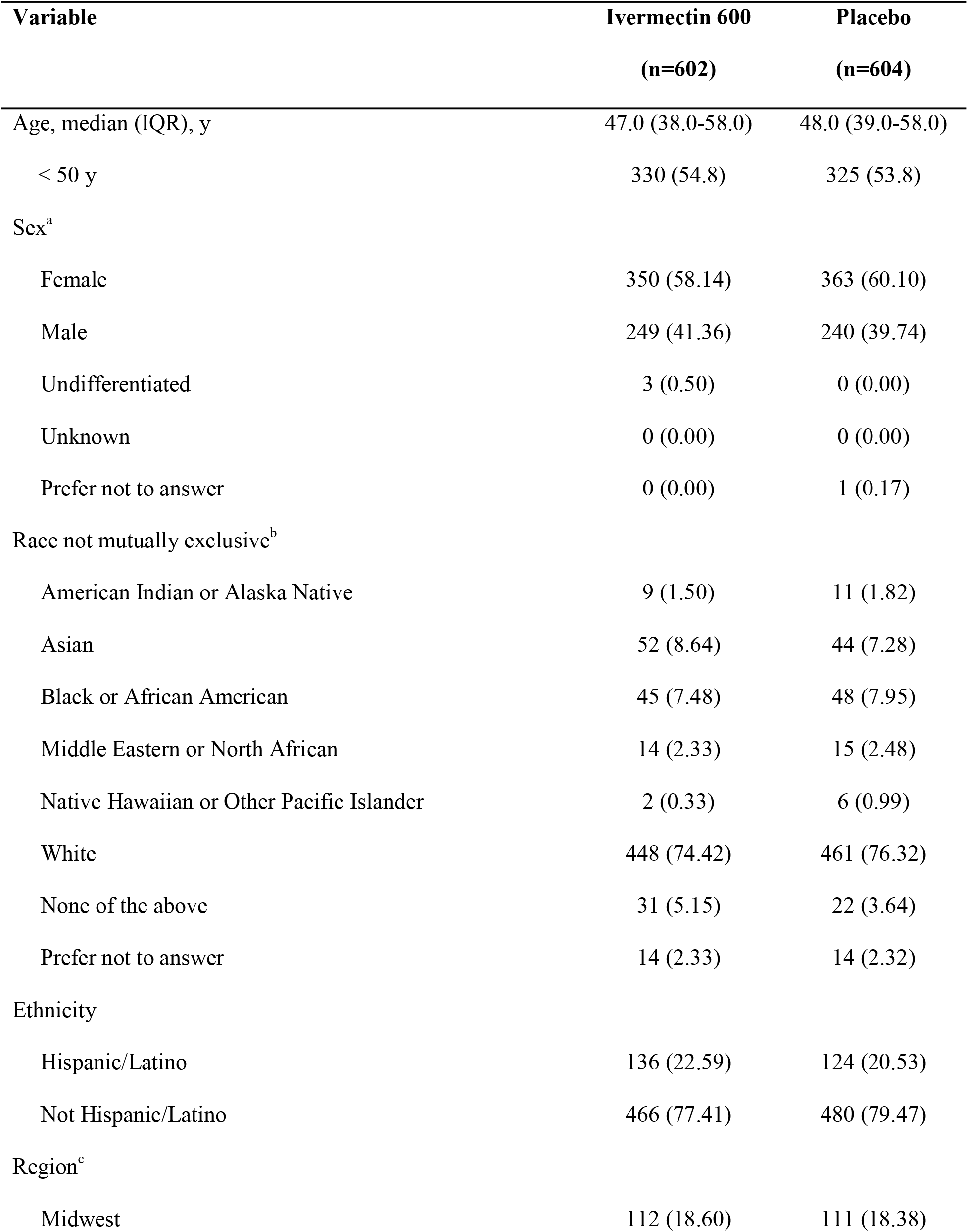
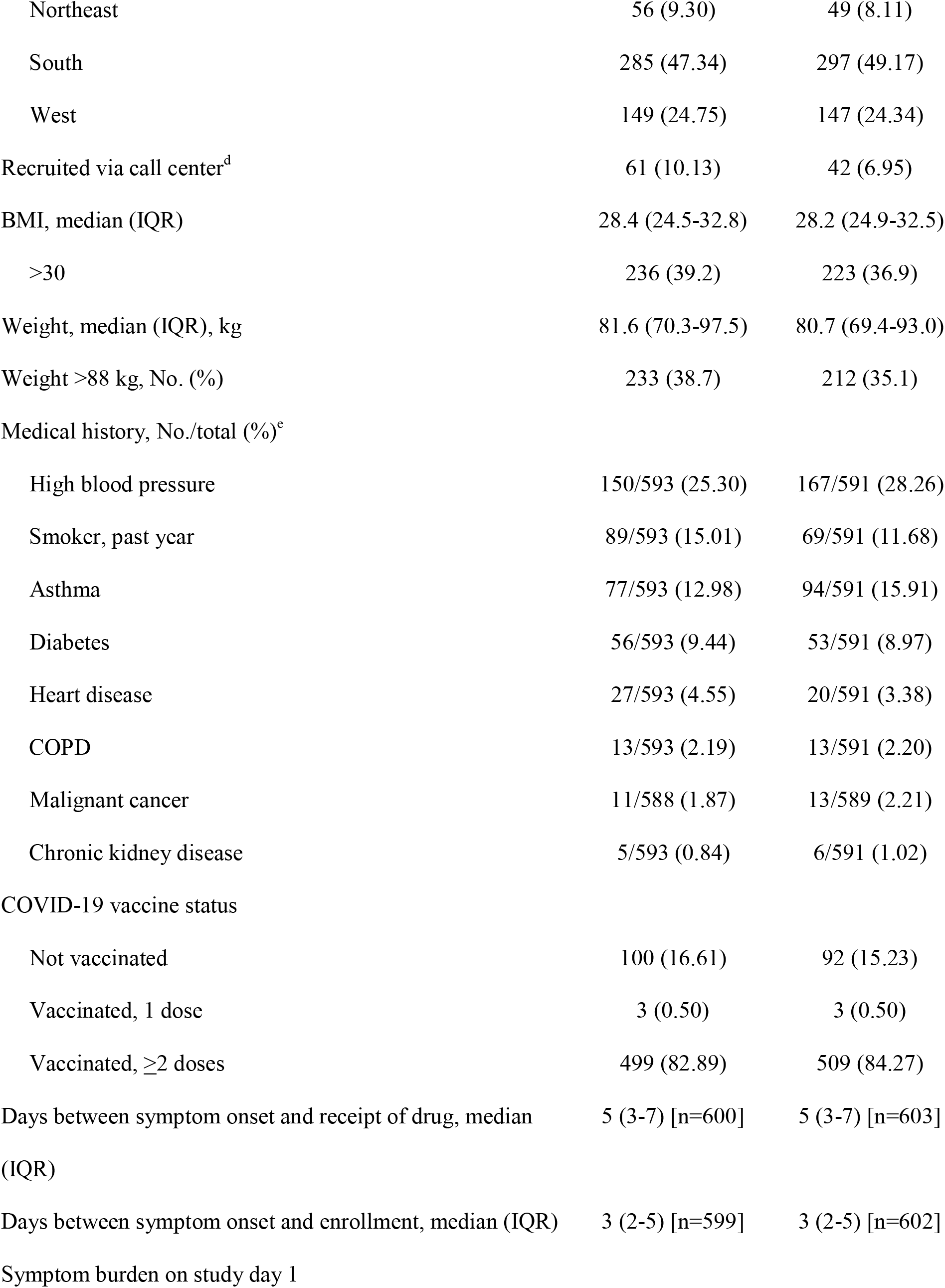
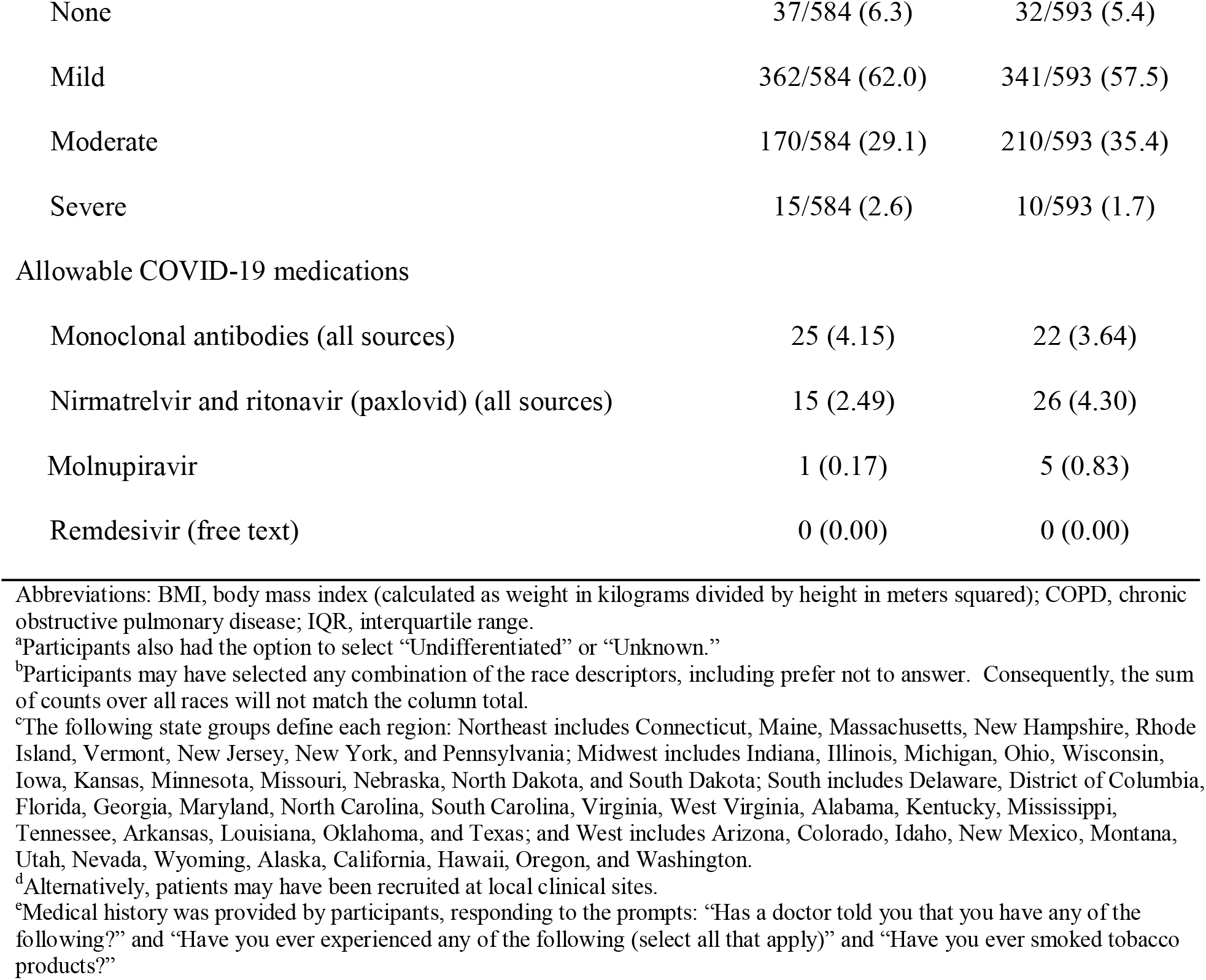
Baseline characteristics

| **Variable** | **Ivermectin 600**<br>**(n=602)** | **Placebo**<br>**(n=604)** |
| --- | --- | --- |
| Age, median (IQR), y | 47.0 (38.0-58.0) | 48.0 (39.0-58.0) |
| < 50 y | 330 (54.8) | 325 (53.8) |
| Sex<sup>a</sup> |  |  |
| Female | 350 (58.14) | 363 (60.10) |
| Male | 249 (41.36) | 240 (39.74) |
| Undifferentiated | 3 (0.50) | 0 (0.00) |
| Unknown | 0 (0.00) | 0 (0.00) |
| Prefer not to answer | 0 (0.00) | 1 (0.17) |
| Race not mutually exclusive<sup>b</sup> |  |  |
| American Indian or Alaska Native | 9 (1.50) | 11 (1.82) |
| Asian | 52 (8.64) | 44 (7.28) |
| Black or African American | 45 (7.48) | 48 (7.95) |
| Middle Eastern or North African | 14 (2.33) | 15 (2.48) |
| Native Hawaiian or Other Pacific Islander | 2 (0.33) | 6 (0.99) |
| White | 448 (74.42) | 461 (76.32) |
| None of the above | 31 (5.15) | 22 (3.64) |
| Prefer not to answer | 14 (2.33) | 14 (2.32) |
| Ethnicity |  |  |
| Hispanic/Latino | 136 (22.59) | 124 (20.53) |
| Not Hispanic/Latino | 466 (77.41) | 480 (79.47) |
| Region<sup>c</sup> |  |  |
| Midwest | 112 (18.60) | 111 (18.38) |
| Northeast | 56 (9.30) | 49 (8.11) |
| South | 285 (47.34) | 297 (49.17) |
| West | 149 (24.75) | 147 (24.34) |
| Recruited via call center<sup>d</sup> | 61 (10.13) | 42 (6.95) |
| BMI, median (IQR) | 28.4 (24.5-32.8) | 28.2 (24.9-32.5) |
| >30 | 236 (39.2) | 223 (36.9) |
| Weight, median (IQR), kg | 81.6 (70.3-97.5) | 80.7 (69.4-93.0) |
| Weight >88 kg, No. (%) | 233 (38.7) | 212 (35.1) |
| Medical history, No./total (%)<sup>e</sup> |  |  |
| High blood pressure | 150/593 (25.30) | 167/591 (28.26) |
| Smoker, past year | 89/593 (15.01) | 69/591 (11.68) |
| Asthma | 77/593 (12.98) | 94/591 (15.91) |
| Diabetes | 56/593 (9.44) | 53/591 (8.97) |
| Heart disease | 27/593 (4.55) | 20/591 (3.38) |
| COPD | 13/593 (2.19) | 13/591 (2.20) |
| Malignant cancer | 11/588 (1.87) | 13/589 (2.21) |
| Chronic kidney disease | 5/593 (0.84) | 6/591 (1.02) |
| COVID-19 vaccine status |  |  |
| Not vaccinated | 100 (16.61) | 92 (15.23) |
| Vaccinated, 1 dose | 3 (0.50) | 3 (0.50) |
| Vaccinated, ≥2 doses | 499 (82.89) | 509 (84.27) |
| Days between symptom onset and receipt of drug, median (IQR) | 5 (3-7) [n=600] | 5 (3-7) [n=603] |
| Days between symptom onset and enrollment, median (IQR) | 3 (2-5) [n=599] | 3 (2-5) [n=602] |
| Symptom burden on study day 1 |  |  |
| None | 37/584 (6.3) | 32/593 (5.4) |
| Mild | 362/584 (62.0) | 341/593 (57.5) |
| Moderate | 170/584 (29.1) | 210/593 (35.4) |
| Severe | 15/584 (2.6) | 10/593 (1.7) |
| Allowable COVID-19 medications |  |  |
| Monoclonal antibodies (all sources) | 25 (4.15) | 22 (3.64) |
| Nirmatrelvir and ritonavir (paxlovid) (all sources) | 15 (2.49) | 26 (4.30) |
| Molnupiravir | 1 (0.17) | 5 (0.83) |
| Remdesivir (free text) | 0 (0.00) | 0 (0.00) |

Abbreviations: BMI, body mass index (calculated as weight in kilograms divided by height in meters squared); COPD, chronic obstructive pulmonary disease; IQR, interquartile range.
<sup>a</sup>Participants also had the option to select “Undifferentiated” or “Unknown.”
<sup>b</sup>Participants may have selected any combination of the race descriptors, including prefer not to answer. Consequently, the sum of counts over all races will not match the column total.
<sup>c</sup>The following state groups define each region: Northeast includes Connecticut, Maine, Massachusetts, New Hampshire, Rhode Island, Vermont, New Jersey, New York, and Pennsylvania; Midwest includes Indiana, Illinois, Michigan, Ohio, Wisconsin, Iowa, Kansas, Minnesota, Missouri, Nebraska, North Dakota, and South Dakota; South includes Delaware, District of Columbia, Florida, Georgia, Maryland, North Carolina, South Carolina, Virginia, West Virginia, Alabama, Kentucky, Mississippi, Tennessee, Arkansas, Louisiana, Oklahoma, and Texas; and West includes Arizona, Colorado, Idaho, New Mexico, Montana, Utah, Nevada, Wyoming, Alaska, California, Hawaii, Oregon, and Washington.
<sup>d</sup>Alternatively, patients may have been recruited at local clinical sites.
<sup>e</sup>Medical history was provided by participants, responding to the prompts: “Has a doctor told you that you have any of the following?” and “Have you ever experienced any of the following (select all that apply)” and “Have you ever smoked tobacco products?”

### Primary Outcome

The median time to recovery was 11 days (IQR 11–12) in the ivermectin group and 11 days (IQR 11–12) in the placebo group. The posterior probability for benefit was 0.68 for the primary outcome of time to recovery with a HR of 1.02 (95% credible interval [CrI], 0.92–1.13) where HR>1 is for faster symptom resolution with ivermectin (**Table 2, Figure 2A**). This posterior probability for the primary outcome was below the prespecified threshold of 0.95 probability (**Supplement 2**). The data do not provide evidence of a treatment benefit even when using a Bayesian non-informative prior, no prior, with various approaches to imputing missing symptom data, or when restricting the analysis to participants who received drug within 2 or 3 days of symptom onset and across severity of symptoms reported on Day 1 (**Table 2, Figure 3, eFigures 3 and 4**). The probability that ivermectin reduced symptom duration by 24 hours was less than 0.1%.

**Figure 2.**
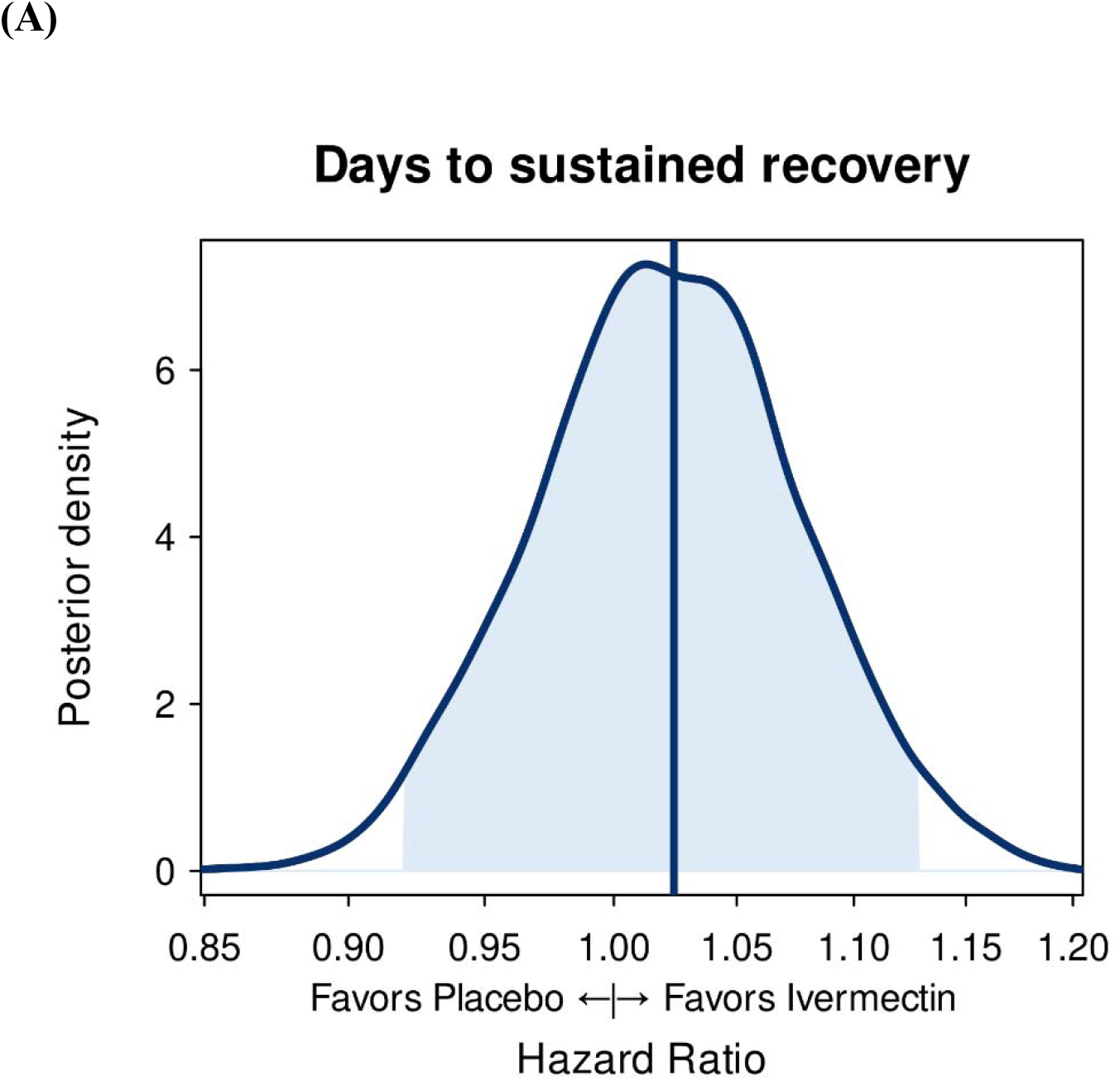
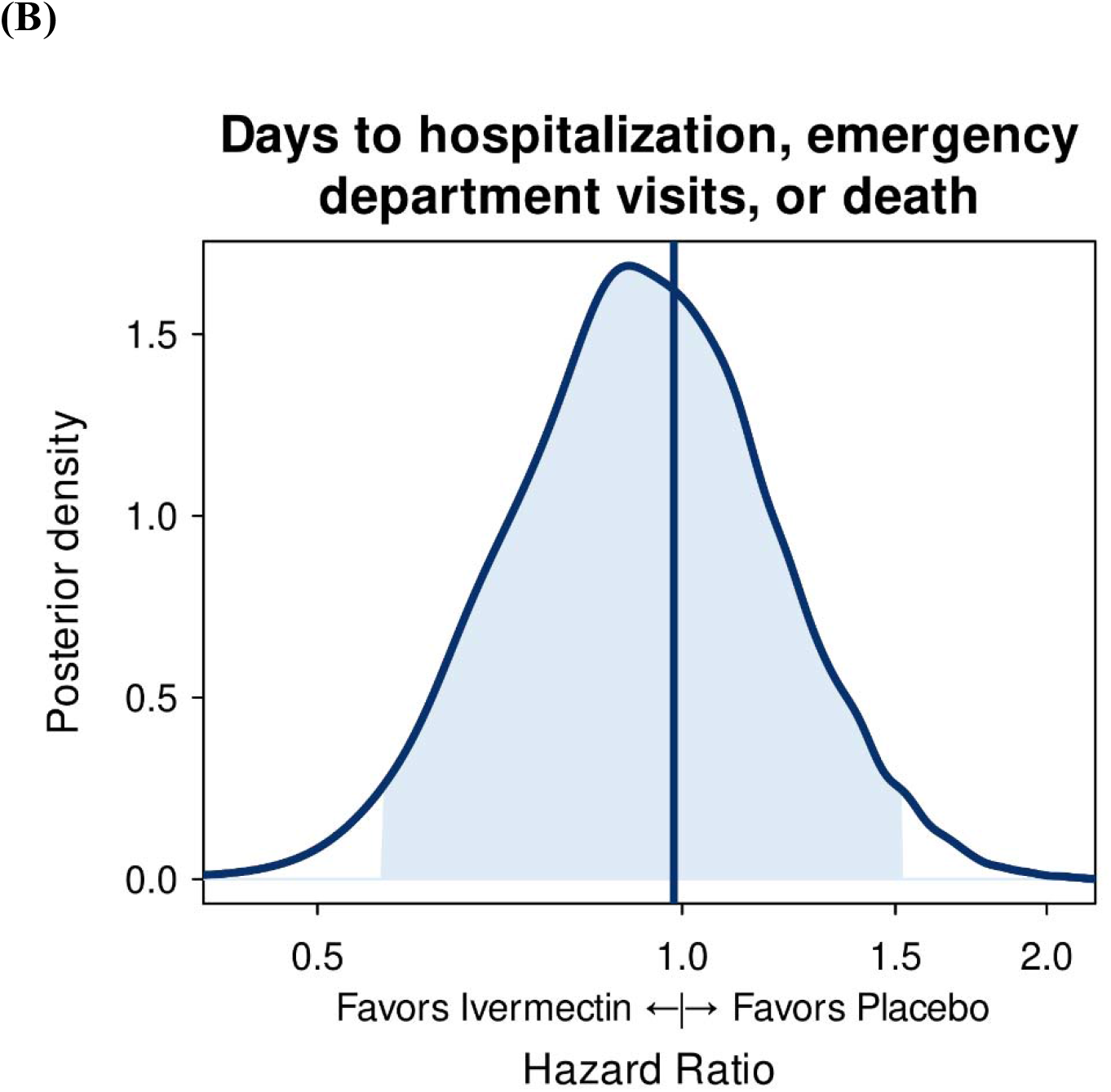
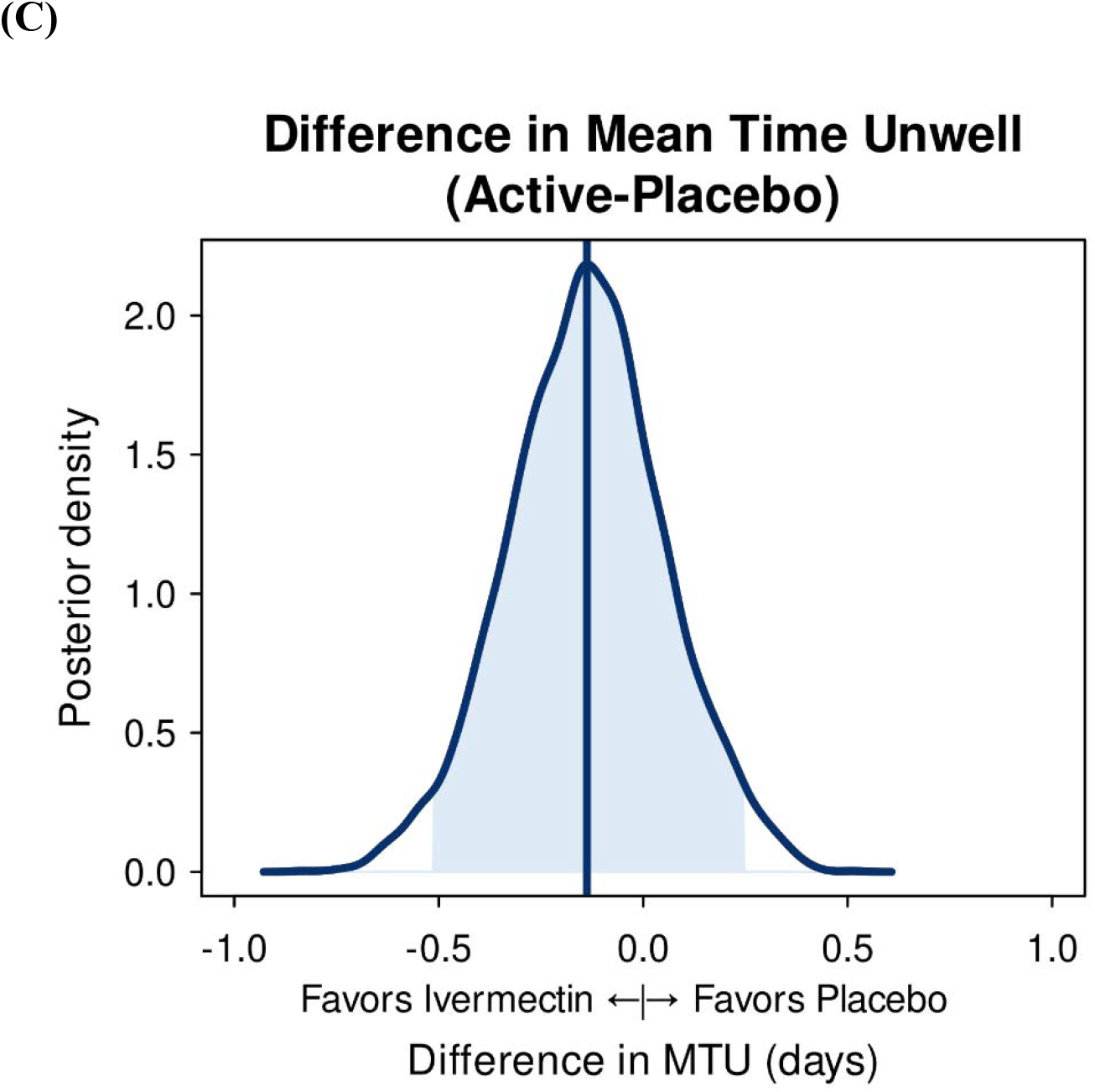
Posterior Distributions of Effects of (A) Time to Sustained Recovery (n=1206); (B) Hospitalization, Urgent or Emergency Care Visits, or Death (n=1206); and (C) Mean Time Unwell (n=1205). Thick vertical lines denote the estimated mean of the posterior distribution. Density is the relative likelihood of posterior probability distribution. Outcomes with higher density are more likely than outcomes with lower density.

**Table 2.** Primary and secondary outcomes

| Endpoint | Ivermectin<br>600<br>(n=602) | Placebo<br>(n=604) | Adjusted estimate<br>(95% CrI)<sup>a</sup> | Posterior<br>P(efficacy) |
| --- | --- | --- | --- | --- |
| **Primary end point, time to recovery<sup>b</sup>** |  |  |  |  |
| Skeptical prior (primary analysis) |  |  | HR, 1.02 (0.92 to 1.13) | 0.68 |
| Skeptical prior (matched/unmatched placebos) |  |  | HR, 1.03 (0.93 to 1.14) | 0.70 |
| Non-informative prior (sensitivity analysis) |  |  | HR, 1.03 (0.91 to 1.17) | 0.69 |
| No prior (sensitivity analysis) |  |  | HR, 1.03 (0.91 to 1.17) | NE<sup>c</sup> |
| **Secondary end points** |  |  |  |  |
| Mortality at day 28 | 1 (0.17) | 0 (0.00) |  |  |
| Hospitalization or death through day 28 | 5 (0.83) | 2 (0.33) | HR, 2.51 (0.49 to 12.96)<sup>c</sup> |  |
| Hospitalization, urgent care, ED visit, or death through day 28 | 34 (5.65) | 36 (5.96) | HR, 1.0 (0.6 to 1.5) | 0.527 |
| **Clinical progression ordinal outcome scale<sup>d</sup>** |  |  |  |  |
| Day 7 |  |  | OR: 1.61 (0.87 to 2.46) | 0.044 |
| No. |  |  | 1206 |  |
| Day 14 |  |  | OR: 2.14 (0.87 to 3.77) | 0.029 |
| No. |  |  | 1175 |  |
| Day 28 |  |  | OR: 2.61 (0.77 to 4.80) | 0.019 |
| No. |  |  | 1206 |  |
| Time unwell, mean (95% CrI), d<sup>e</sup> | 11.21 (11.01 to 11.41) | 11.35 (11.16 to 11.54) | $\Delta$: -0.14 (-0.51 to 0.24) | 0.772 |
| Days benefit, mean (95%CrI), d | 3.42 (3.18 to 3.64) | 3.26 (3.03 to 3.48) | $\Delta$: 0.16 (-0.28 to 0.61) | 0.772 |

Abbreviations: ED, emergency department; HR, hazard ratio; OR, odds ratio; NE, not estimated
<sup>a</sup> Unless otherwise noted, a highest-density credible interval. Adjustment variables for time to recovery, mortality, composite clinical endpoints, and clinical progression in addition to randomization assignment: age (as restricted cubic spline), sex, duration of symptoms prior to receipt of study drug, calendar time (as restricted cubic spline), vaccination status, geographic region (Northeast, Midwest, South, West), call center indicator, and baseline symptom severity. For time to recovery, HR > 1 is favorable for faster recovery for ivermectin compared with placebo. For the secondary endpoints, HR < 1, OR < 1, and $\Delta$ < 0 is favorable for ivermectin.
<sup>b</sup> Time to recovery is from receipt of study drug to achieving the third of 3 days of recovery. HR > 1.0 is favorable for faster recovery for ivermectin compared with placebo
<sup>c</sup> Confidence interval, low event rate precluded covariate adjustment
<sup>d</sup> The description of the 8 levels of the clinical progression ordinal outcome scale is reported in the supplement. Proportional odds was not evaluated as the vast majority of participants were either at home with limitations or at home without limitations, resulting in a model that is approximately a logistic regression.
<sup>e</sup> Adjustment variables for mean time unwell in addition to randomization assignment: age and calendar time.

**Figure 3.**
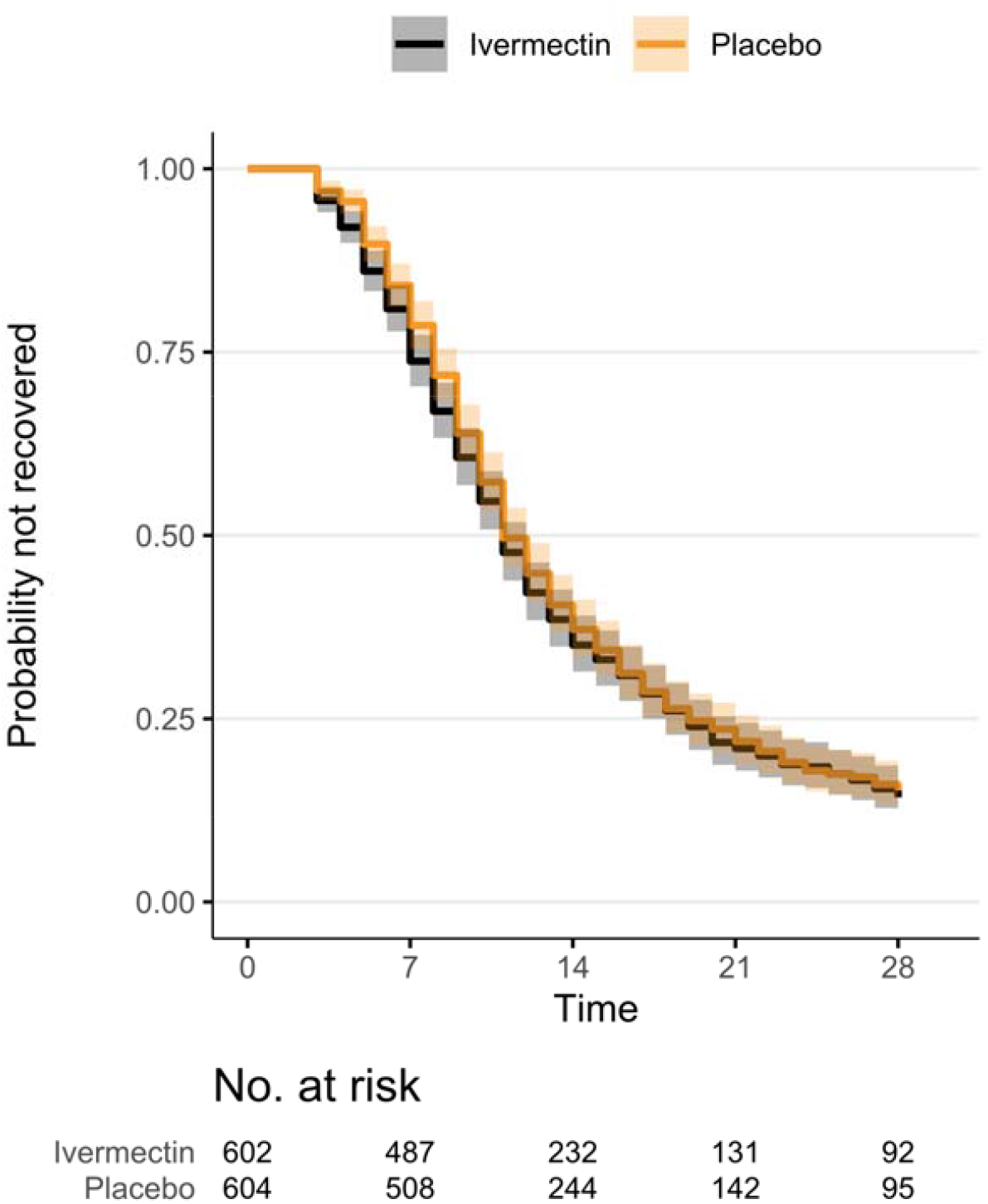
Kaplan-Meier for Primary Outcome of Time to Sustained Recovery. Recovery occurs on the third of 3 consecutive days without symptoms. Four participants were censored for nonresponse, and all others were followed up until recovery, death, or the end of short-term 28-day follow-up. Median (IQR) time to recovery was 11 days (11, 12) in the ivermectin group and 11 days (11, 12) in the placebo group.

### Secondary Outcomes

Hospitalizations and deaths were uncommon, with 5 events (1 death not attributable to Covid-19 or treatment) in the ivermectin group and 2 events (no deaths) in the placebo group (**eFigure 5A**). Statistical comparisons were not done as they would be uninformative with so few events. The composite secondary outcome of urgent care or ED visits, hospitalizations, or death was not shown to differ with ivermectin (5.6% [34/602]) compared with placebo (6.0% [36/604]) (HR 1.0, 95% CrI 0.6–1.5, P(HR<1=0.53) (**Table 2, Figure 2B, eFigure 5B**). The difference in the amount of time spent feeling unwell with COVID-19 was estimated as 3 hours and 20 minutes faster with ivermectin (95% CrI, 12 hours better to 6 hours worse) than placebo (**Figure 2C**). The COVID Clinical Progression Scale at days 7, 14, and 28 did not meet prespecified thresholds for beneficial treatment effect (**Supplement 3**). For example, by day 7, 88% (532/602) of the ivermectin group and 91% (549/604) of the placebo group were not hospitalized and did not report limitation of activities (**eFigure 6**).

### Heterogeneity of Treatment Effect Analyses

Interaction tests for heterogeneity of treatment effect showed no overall influence of the putative treatment effect modifiers even when all subgroup analyses across symptom severity were not adjusted for multiple comparisons (**eFigure 7**). The overall effect of timing from symptom onset to receipt of study drug was not significant (p=0.15 for heterogeneity). Similarly, no evidence existed for a different treatment effect of ivermectin compared with placebo for severity of symptoms, sex, age, body mass index, calendar time, or vaccination status.

### Adverse Events

Among participants who reported taking study drug at least once, adverse events were similar in both groups (9.2% [52/566] with ivermectin and 7.1% [41/576] with placebo) (**eTable 2**). Adverse events reported >2 times only in the ivermectin group included cognitive impairment (n=4), blurred vision (n=5), light sensitivity to eye (n=5), photophobia (n=4), dizziness (n=5), and asthma (n=3). Serious adverse events were rare: 5 with ivermectin and 3 with placebo. The death in the ivermectin arm was accidental.

## DISCUSSION

Among a largely vaccinated outpatient population with COVID-19, treatment with ivermectin, with a targeted maximum dose of 600 μg/kg, daily for 6 days compared with placebo was not shown to improve time to recovery in over 1200 participants in the US during omicron variant/subvariant circulation. No evidence of benefit was observed for secondary clinical outcomes including the composite of hospitalization, death, or acute care visits. Hospitalization and death were uncommon in this largely vaccinated population. These findings do not support the use of ivermectin in outpatients with COVID-19.

Multiple large, double-blind randomized controlled trials have failed to identify a clinically meaningful benefit of ivermectin when used at a targeted dose of 400 μg/kg daily for 3 days.^6,16^ This large clinical trial addresses a potential gap in knowledge by testing 1) a higher daily dose (targeted maximum dose of 600 μg/kg) and 2) longer 6-day duration of ivermectin,. Due to the lack of early phase studies or animal model studies to determine optimal dosing for a therapeutic drug, the appropriate dosing of ivermectin for COVID-19 was never determined. A combination of modeling studies and a proof-of-concept clinical study have suggested doses up to 600 μg/kg daily may achieve system levels sufficient for *in vitro* antiviral activity.^17,18^ Although a phase 2 trial testing ivermectin 600 μg/kg daily for 7 days and assessing a virologic endpoint of oropharyngeal SARS-CoV-2 PCR did not show measurable antiviral activity and was stopped for futility.^21^ This dose was safe and generally well tolerated, with a higher prevalence of the known self-resolving visual disturbances in the intervention arm previously reported with similar doses of ivermectin for parasitic infections.^17,18^

Recent results from the current platform trial reported participants receiving ivermectin, with a 400 μg/kg, daily for 3 days had a an average ∼12-hour shorter time spent unwell when compared with placebo, a secondary outcome for the trial.^15^ This finding was not replicated here. The notable difference in baseline characteristics between these 2 cohorts is the completed vaccination rate, which is 84% for this study and was 47% for the prior ivermectin 400 μg/kg group.^15^ Hospitalizations and COVID-19-related clinical events were less common in this largely vaccinated cohort. The incidence of acute care visits, hospitalizations, or death was similar with ivermectin (5.7%) and placebo (6.0%); a result also observed in the two prior ivermectin 400 μg/kg randomized trials in the US.^6,15^

This trial has several strengths. This was a double-blind, randomized, placebo-controlled nationwide trial with 93 enrolling sites and a call center that recruited participants from all 50 US states. The ivermectin 600 μg/kg group of the platform trial enrolled rapidly due to ongoing omicron variant/subvariant surges and largely included vaccinated people, thus representing a highly relevant study population that also addresses a weakness of many other studies that excluded vaccinated people.

### Limitations

This study has limitations. Due to infrequent hospitalization, we cannot draw definitive inferences on whether ivermectin impacts hospitalization without much larger trials. Also, due to the remote nature of the trial, 60% of participants received study drug within 5 days of symptom onset. Most outpatient COVID-19 antiviral trials have limited enrollment to participants within 5 days of symptom onset.^1,2^ In this trial, we observed no evidence of a differential treatment effect based on shorter time to study drug receipt.

## Conclusions

Among outpatients with mild or moderate COVID-19, treatment with ivermectin, with a targeted maximum dose of 600 μg/kg daily for 6 days, was not shown to improve time to recovery compared with placebo. These findings do not support the use of ivermectin in outpatients with COVID-19.

## Data Availability

ACTIV-6 is a platform trial using shared placebos. On completion of the platform trial when there is no risk of unblinding across study arms the data will be made publicly available by depositing it in an approved data repository such as NHLBI BioData Catalyst.

## Acknowledgments

We thank Samuel Bozzette, MD, PhD, and Eugene Passamani, MD, both of the National Center for Advancing Translational Sciences (NCATS), for their roles in the trial design and protocol development. We also thank the ACTIV-6 Data Monitoring Committee and Clinical Events Committee Members (listed below) for their contributions.

Data Monitoring Committee: Clyde Yancy, MD, MSc, Northwestern University Feinberg School of Medicine; Adaora Adimora, MD, University of North Carolina, Chapel Hill; Susan Ellenberg, PhD, University of Pennsylvania; Kaleab Abebe, PhD, University of Pittsburgh; Arthur Kim, MD, Massachusetts General Hospital; John D. Lantos, MD, Children’s Mercy Hospital; Jennifer Silvey-Cason, Participant representative; Frank Rockhold, PhD, Duke Clinical Research Institute; Sean O’Brien, PhD, Duke Clinical Research Institute; Frank Harrell, PhD, Vanderbilt University Medical Center; Zhen Huang, MS, Duke Clinical Research Institute.

Clinical Events Committee: Renato Lopes, MD, PhD, MHS, W. Schuyler Jones, MD, Antonio Gutierrez, MD, Robert Harrison, MD, David Kong, MD, Robert McGarrah, MD, Michelle Kelsey, MD, Konstantin Krychtiuk, MD, Vishal Rao, MD all of the Duke Clinical Research Institute, Duke University School of Medicine.

## Author Contributions

Drs Naggie and Hernandez had full access to all of the blinded data in the study. Drs Lindsell and Stewart directly accessed and verified the underlying study data and take responsibility for the integrity of the data and the accuracy of the data analysis. All authors contributed to the drafting and review of the manuscript and agreed to submit for publication.

## Data Sharing Statement

Available in the Supplement to the ACTIV-6 Statistical Analysis Plan (**Supplement 4**).

